# Characterising the Stability of Polygenic Risk Scores: implications for risk stratification

**DOI:** 10.64898/2026.05.17.26353273

**Authors:** Andreza Ferreira, Penelope A. Lind, Hayley Moody, Ian B. Hickie, Catherine M. Olsen, David C. Whiteman, Matthew H. Law, Dan J. Siskind, Nicholas G. Martin, Richard C. Medland, Sarah E. Medland

## Abstract

Polygenic risk scores (PRS) improve progressively as genome-wide association studies (GWAS) increase in sample size and ancestral diversity, yet the effect of successive GWAS releases on individual PRS rankings remains poorly characterised. Here, we quantify how individual PRS rankings change across GWAS releases, whether those changes favour cases over controls, how consistently individuals maintain their relative position, and whether those in high-risk strata retain that classification over time. Using PRS derived from four GWAS releases for bipolar disorder, major depressive disorder, and schizophrenia in three Australian cohorts, we observed widespread bidirectional reclassification that exceeded the theoretical minimum of expected reclassification, and was directionally consistent with case-control status when discriminative performance improved. Rank variability was substantial and uniformly distributed across all levels of risk, rank persistence was limited across releases, and retention of high-risk classifications was variable across disorders and largely accounted for by the inter-release correlation. These findings demonstrate that individual PRS rankings are dynamic and shaped by progressive improvements in effect-size estimates, carrying important implications for PRS-based risk stratification strategies that rely on stable classifications in psychiatric research and clinical practice.

## Main

Psychiatric disorders, including bipolar disorder (BD), major depressive disorder (MDD), and schizophrenia (SCZ), are complex and highly heritable phenotypes whose polygenic architecture has been progressively characterised through genome-wide association studies (GWAS)^1,2,3^. The genetic contribution to these phenotypes is distributed across numerous variants of modest effect, and successive GWAS iterations, driven by expanding discovery samples and improved effect-size estimation, have broadened the set of identified loci and increased the proportion of heritability explained^4,5,6^. These advances have consistently improved the discriminative performance of polygenic risk scores (PRS) for estimating individual genetic liability using GWAS-derived weights. As a result, PRS are now widely used in research and risk-stratification contexts, where genetic risk estimates may inform participant selection and guide personalised treatment strategies^7,8,9^.

As effect-size estimates converge towards their population values with increasing statistical power, PRS derived from updated summary statistics provide more accurate estimates of individual polygenic liability^10,11^. This process leads to the reclassification of individuals along the population risk distribution, reflecting the increasing concordance between observed rankings and underlying genetic liability^12,13^. Quantifying this reclassification is relevant to the design of longitudinal studies^14^, the interpretation of PRS derived from successive GWAS iterations^15^, and the development of risk-stratification strategies that incorporate ongoing improvements in genetic prediction^16,17^. Although prior work has documented variability in PRS rankings across scores derived from different discovery samples, methods, or platforms^18,19,20,21,22^, the distributional properties of reclassification induced by successive GWAS releases remain comparatively unexplored. Further research is needed to characterise: (i) how individual PRS rankings change across successive GWAS releases, and the extent to which these changes exceed what would be expected from improvements in discriminative performance alone; (ii) whether such changes are directionally consistent with case-control status; (iii) how consistently individuals preserve their relative position across releases; and (iv) the extent to which individuals in high-risk strata retain their classification.

In this study, we evaluated how individual PRS rankings change as effect-size estimates become progressively refined across successive GWAS releases, using data from three independent cohorts: the Genetics of Bipolar Disorder Study^23^ (GBP; *n* = 3,039), the Australian Genetics of Depression Study^24^ (AGDS; *n* = 13,659), and the ClozaGene Study^25^ (CLZ; *n* = 765), with a shared control sample drawn from the QSkin Sun and Health Study^26^ (QSkin2; *n* = 7,320). PRS were constructed using SBayesRC^27^ applied to summary statistics from four successive GWAS releases for BD, MDD, and SCZ from the Psychiatric Genomics Consortium (PGC) (Methods). We quantified changes in individual PRS rankings across successive releases as a measurable consequence of iterative effect-size re-estimation, and characterised this process along four complementary dimensions: the magnitude of reclassification, its directional consistency with case-control status, within-individual rank variability and persistence, and retention within high-risk classifications. To distinguish reclassification attributable to genuine gains in discriminative performance from reclassification arising independently of those gains, we derived a theoretical minimum expected reclassification from changes in *R*^2^ and inter-release correlation, which served as a benchmark for empirically observed reclassification. These findings establish PRS rank variability as a structural dimension of polygenic prediction and provide a quantitative foundation for understanding how advances in psychiatric genomics progressively refine the definition of individual risk.

## Results

### GWAS discovery sample size and discriminative performance increase across successive releases

Successive GWAS releases do not merely improve predictive performance; they progressively revise the effect-size estimates from which individual PRS rankings are derived. We first contextualised changes in individual PRS rankings relative to this evolving discovery landscape across successive GWAS releases for BD, MDD, and SCZ in three Australian cohorts (Methods). Across all three disorders, increases in discovery sample size, the number of associated loci, and discriminative performance were substantial (Table 1). Discriminative performance improved across most successive GWAS releases. For BD, pseudo-*R*^2^ increased from 0.021 (AUC = 0.572) in 2011 to 0.061 (AUC = 0.629) in 2024; for MDD, from 0.003 (AUC = 0.529) in 2013 to 0.034 (AUC = 0.594) in 2025; and for SCZ, from 0.147 (AUC = 0.744) in 2011 to a peak of 0.300 (AUC = 0.843) in the 2014 release, representing the highest discriminative performance observed across all disorders and releases, consistent with the stronger polygenic architecture of SCZ. The most recent SCZ release was associated with a modest reduction in discriminative performance relative to the preceding release.

**Table 1:** GWAS discovery sample characteristics and discriminative performance of derived PRS.

| Disorder | Year | Cases | Controls | Total N | $h^2_{\text{SNP}}$ | GW loci | Pseudo $R^2$ | $\beta_{\text{PRS}}$ [95% CI] | $p$-value | AUC |
| --- | --- | --- | --- | --- | --- | --- | --- | --- | --- | --- |
| BD | 2011 | 11,974 | 51,792 | 63,766 | * | 2 | 0.021 | 0.70 [0.59–0.82] | $2.28 \times 10^{-34}$ | 0.572 |
| | 2019 | 29,764 | 169,118 | 198,882 | 0.17–0.23 | 30 | 0.045 | 0.64 [0.57–0.71] | $1.33 \times 10^{-70}$ | 0.608 |
| | 2021 | 41,917 | 371,549 | 413,466 | 0.186 | 64 | 0.061 | 0.70 [0.63–0.76] | $1.17 \times 10^{-94}$ | 0.627 |
| | 2024 | 40,992<sup>a</sup> | 80,215<sup>a</sup> | 121,207<sup>a</sup> | 0.22 | 298 | 0.061 | 0.87 [0.79–0.95] | $4.69 \times 10^{-95}$ | 0.629 |
| MDD | 2013 | 9,240 | 9,519 | 18,759 | * | 0 | 0.003 | 1.80 [1.30–2.31] | $1.95 \times 10^{-12}$ | 0.529 |
| | 2018 | 135,458 | 344,901 | 480,359 | 0.087 | 44 | 0.015 | 0.82 [0.71–0.93] | $4.42 \times 10^{-50}$ | 0.562 |
| | 2019 | 246,363 | 561,190 | 807,553 | 0.089 | 102 | 0.018 | 1.42 [1.25–1.59] | $6.04 \times 10^{-61}$ | 0.568 |
| | 2025 | 525,197<sup>a</sup> | 3,362,335<sup>a</sup> | 3,887,532<sup>a</sup> | 0.084 | 697 | 0.034 | 1.03 [0.94–1.12] | $1.86 \times 10^{-110}$ | 0.594 |
| SCZ | 2011 | 17,836 | 33,859 | 51,695 | * | 7 | 0.147 | 1.49 [1.36–1.62] | $6.82 \times 10^{-111}$ | 0.744 |
| | 2013 | 21,246 | 52,129 | 73,375 | 0.32–0.33 | 22 | 0.214 | 1.42 [1.31–1.53] | $6.59 \times 10^{-147}$ | 0.794 |
| | 2014 | 32,405<sup>a</sup> | 42,221<sup>a</sup> | 74,626<sup>a</sup> | 0.23 | 108 | 0.300 | 1.41 [1.31–1.50] | $2.01 \times 10^{-187}$ | 0.843 |
| | 2022 | 67,390<sup>a</sup> | 94,015<sup>a</sup> | 161,405<sup>a</sup> | 0.24 | 287 | 0.245<sup>b</sup> | 1.31 [1.22–1.40] | $1.11 \times 10^{-164}$ | 0.815<sup>b</sup> |

BD, bipolar disorder; MDD, major depressive disorder; SCZ, schizophrenia. $h^2_{\text{SNP}}$ , SNP-based heritability estimated from the GWAS discovery sample; GW loci, number of independent genome-wide significant loci ( $p < 5 \times 10^{-8}$ ) reported in the original publication. Pseudo $R^2$ , Nagelkerke pseudo- $R^2$ from a logistic regression of case-control status on the PRS; $\beta_{\text{PRS}}$ , log-odds regression coefficient for the standardised PRS [95% CI]; $p$ -value, Wald test $p$ -value for $\beta_{\text{PRS}}$ ; AUC, area under the receiver-operating characteristic curve. <sup>a</sup> For multi-ancestry releases, cases and controls reflect the EUR-ancestry subset, and the number of genome-wide significant loci reflects the full multi-ancestry meta-analysis; polygenic risk scores were derived from the corresponding EUR-ancestry summary statistics. <sup>b</sup> Discriminative performance was attenuated relative to the preceding release, coinciding with the transition to multi-ancestry discovery designs.

### Empirical reclassification exceeds the theoretical minimum and is directionally consistent with case-control status

Any gain in *R*^2^ has direct implications for individual risk ranking, as improved predictive accuracy necessarily induces some degree of reclassification among individuals whose prior positions were estimated with uncertainty. To quantify the minimum reclassification expected from such gains, we derived the theoretical minimum of expected reclassification under a bivariate normal model (Methods), and compared it with empirical reclassification across successive releases (Table 2; Table 3). Two clear patterns emerged. First, reclassification was directionally consistent with case-control status: whenever discriminative performance improved, cases were preferentially reclassified into higher-risk strata and controls into lower-risk strata at each update. For BD, empirical NRI totals of 0.19 and 0.17 were observed in the two transitions associated with improved performance, with no directional signal observed in the 2021-2024 transition. For MDD, empirical NRI totals were 0.15, 0.03, and 0.13 across successive transitions, with the largest empirical reclassification observed in the 2013-2018 transition, whereas the largest gain in *R*^2^ occurred in the 2019-2025 transition. For SCZ, NRI totals of 0.59 and 0.57 were observed in the first two transitions, with 65-66% of cases reclassified upward and 64% of controls downward, before reversing to −0.46 in the 2014-2022 transition, where discriminative performance declined. Second, the magnitude of empirical reclassification consistently exceeded the theoretical minimum across all improving transitions for all three disorders, with observed-to-expected ratios ranging from 1.59 to 1.93 (Table 3), indicating that individuals move more across the risk distribution than model improvement alone would predict. Comparable excess was evident in the opposite direction for the declining SCZ transition (2014-2022; ratio = 1.69), suggesting that this structural variability operates independently of whether discriminative performance improves or declines. This excess likely reflects not only the correction of imprecise prior rankings, but also the propagation of sampling variance inherent to effect-size re-estimation across successive GWAS releases.

**Table 2:**
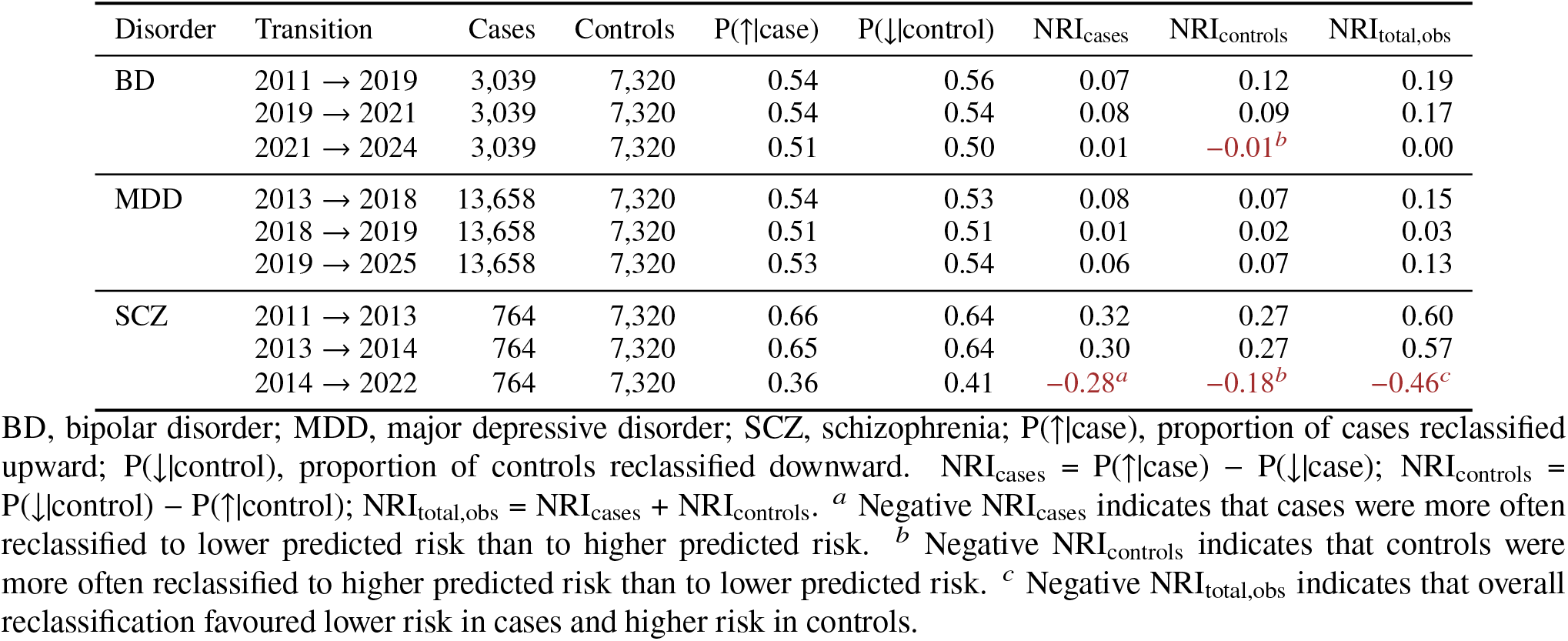
Empirical reclassification across successive GWAS releases.

**Table 3:** Theoretical minimum of expected reclassification derived from Δ*R*^2^ across successive GWAS releases.

| Disorder | Transition | $R^2(t_1)$ | $R^2(t_2)$ | $\Delta R^2$ | $\rho_S$ | $\text{NRI}_{\text{cases}}$ | $\text{NRI}_{\text{controls}}$ | $\text{NRI}_{\text{total,th}}$ | $\vert \text{NRI}_{\text{obs}}\vert /\vert \text{NRI}_{\text{th}}\vert $ |
| --- | --- | --- | --- | --- | --- | --- | --- | --- | --- |
| BD | 2011 → 2019 | 0.021 | 0.045 | +0.024 | 0.56 | 0.06 | 0.06 | 0.12 | 1.59 |
| | 2019 → 2021 | 0.045 | 0.061 | +0.016 | 0.79 | 0.05 | 0.05 | 0.09 | 1.84 |
| | 2021 → 2024 | 0.061 | 0.061 | +0.000 | 0.88 | 0.00 | 0.00 | 0.00 | – |
| MDD | 2013 → 2018 | 0.003 | 0.015 | +0.012 | 0.29 | 0.05 | 0.05 | 0.09 | 1.59 |
| | 2018 → 2019 | 0.015 | 0.018 | +0.003 | 0.42 | 0.01 | 0.01 | 0.02 | 1.93 |
| | 2019 → 2025 | 0.018 | 0.034 | +0.016 | 0.42 | 0.04 | 0.04 | 0.08 | 1.66 |
| SCZ | 2011 → 2013 | 0.147 | 0.214 | +0.067 | 0.86 | 0.16 | 0.16 | 0.32 | 1.88 |
| | 2013 → 2014 | 0.214 | 0.300 | +0.086 | 0.79 | 0.16 | 0.16 | 0.32 | 1.76 |
| | 2014 → 2022 | 0.300 | 0.245 | –0.055 | 0.88 | –0.14<sup>a</sup> | –0.14<sup>a</sup> | –0.27<sup>a</sup> | 1.69<sup>b</sup> |

BD, bipolar disorder; MDD, major depressive disorder; SCZ, schizophrenia; $R^2(t_1)$ , $R^2(t_2)$ , Nagelkerke pseudo- $R^2$ at the earlier ( $t_1$ ) and later ( $t_2$ ) GWAS release (Table 1); $\Delta R^2 = R^2(t_2) - R^2(t_1)$ ; $\rho_S$ , Spearman correlation of polygenic risk score percentile ranks between releases (Table S1); $\text{NRI}_{\text{total,th}}$ , theoretical minimum of expected reclassification from $\Delta R^2$ and $\rho_S$ under a bivariate normal model; $|\text{NRI}_{\text{obs}}|/|\text{NRI}_{\text{th}}|$ , fold-excess of empirical over theoretical reclassification magnitude. <sup>a</sup> Negative $\text{NRI}_{\text{cases}}$ , $\text{NRI}_{\text{controls}}$ , and $\text{NRI}_{\text{total,th}}$ indicate that $\Delta R^2 < 0$ ; interpretation analogous to Table 2, with values reflecting model-predicted rather than observed reclassification. <sup>b</sup> For the declining SCZ transition (2014–2022), the ratio reflects the fold-excess of empirical over theoretical reclassification magnitude, with both quantities in the negative direction. For BD 2021–2024, the ratio is undefined as the theoretical minimum is zero ( $\Delta R^2 = 0$ ).

### Within-individual rank variability is widespread and bidirectional

Individual trajectories of PRS percentile rank across successive GWAS releases were heterogeneous and largely non-monotonic, with many individuals exhibiting substantial fluctuations across the risk distribution, irrespective of initial risk stratum (Figure 1a). When individuals were ordered by their rank at the most recent release, ranks from prior releases appeared substantially less structured across the same individuals, indicating limited rank persistence across successive releases (Figure 1b). To quantify within-individual rank variability across GWAS releases, we computed the coefficient of variation of individual percentile ranks, providing a normalised measure of dispersion relative to average rank (Figure 1c, left). Median CVs indicated substantial rank variability across all disorders, ranging from 28.31% in SCZ to 48.29% in MDD. Upper-tail values revealed even greater variability in a meaningful proportion of individuals: at the 99th percentile, CVs reached 145.52% for BD, 142.54% for MDD, and 125.53% for SCZ, indicating that for some individuals, rank fluctuations across releases exceeded their average percentile position. To distinguish systematic directional drift from bidirectional reclassification, we modelled individual percentile rank as a linear function of release order (Figure 1c, right). Slope distributions were centred near zero across all disorders, with median values of −0.012 (IQR −1.600 to 1.608) for BD, 0.002 (IQR −2.032 to 2.075) for MDD, and 0.030 (IQR −1.174 to 1.178) for SCZ, indicating that successive updates reflect bidirectional reclassification across the risk distribution rather than systematic directional drift.

**Figure 1:**
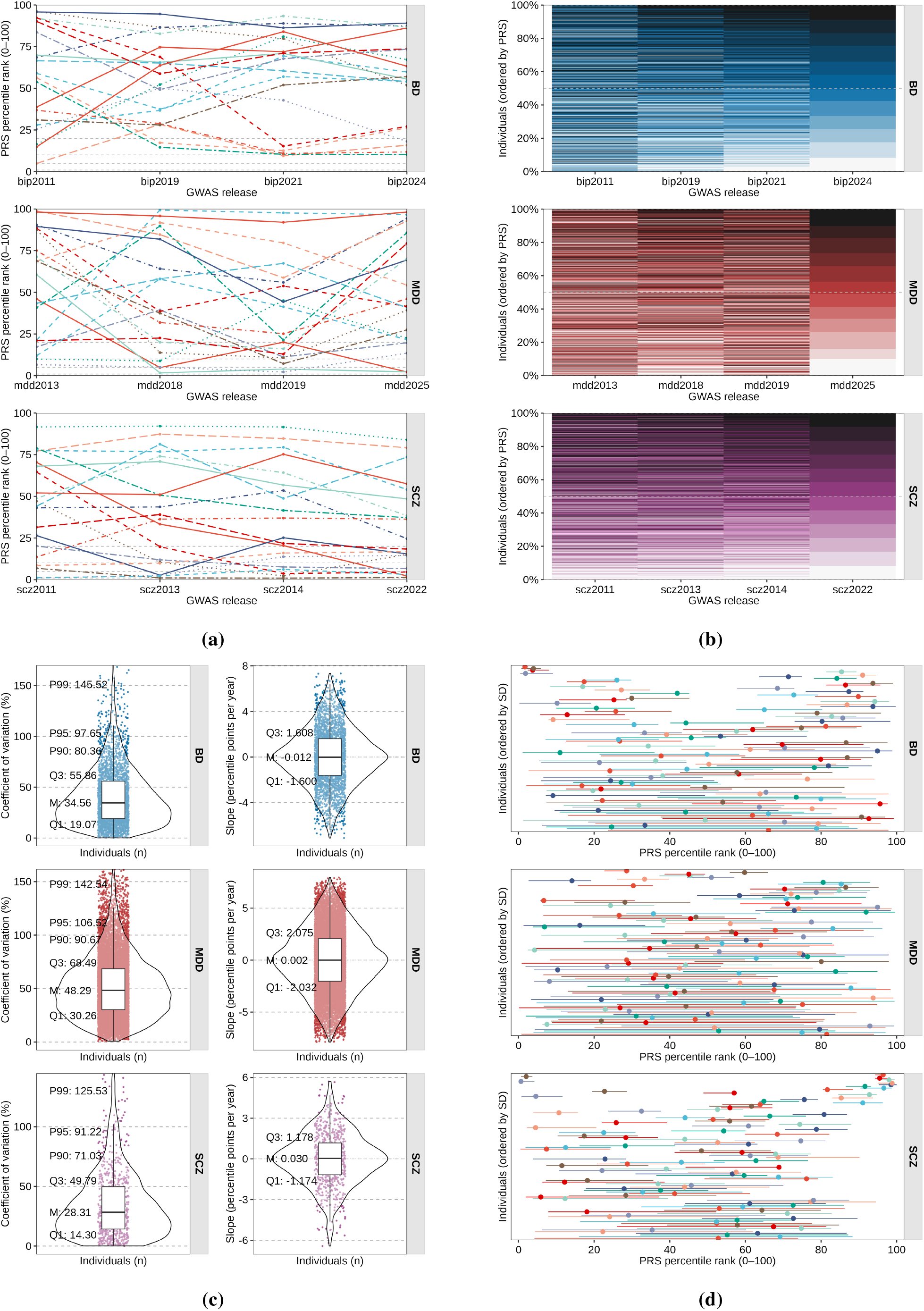
Within-individual variability of PRS percentile ranks across successive GWAS releases. **(a)** Trajectories of PRS percentile ranks for 20 randomly sampled individuals per disorder across successive GWAS releases. **(b)** Heatmap of PRS percentile ranks for 765 randomly sampled individuals per disorder, ordered by rank at the most recent release. **(c)** Violin plots with embedded box plots showing the distribution of the coefficient of variation (CV; left) and linear trend slope (right) of individual PRS percentile ranks across releases. **(d)** Range plots for 100 randomly sampled individuals per disorder, ordered by within-individual standard deviation.

We next examined whether within-individual rank variability was uniformly distributed across the risk distribution or concentrated within particular strata, and quantified the absolute magnitude. Absolute within-individual rank variability was quantified using the standard deviation of percentile ranks across releases, SD(*P*_*i*_) (Table S2; Figure 1d). Rank variability was substantial and uniformly distributed across risk strata. In BD, 27% of individuals had SD(*P*_*i*_) ≥ 20 percentile points, with nearly identical proportions across the bottom 10% (29%), lower-middle (25%), upper-middle (27%), and top 10% (28%) strata, indicating that variability is not concentrated near classification thresholds but is instead a widespread feature of the entire risk distribution. The same pattern was observed in MDD, where 56% of individuals overall had SD(*P*_*i*_) ≥ 20 percentile points, with proportions ranging from 55% in the bottom 10% to 56% in the upper-middle stratum, consistent with the same broadly uniform distribution across strata. Schizophrenia showed comparatively greater rank stability, with only 12% of individuals overall exhibiting SD(*P*_*i*_) ≥ 20 percentile points and 17% exhibiting SD(*P*_*i*_) ≤ 5 percentile points. Rank variability also remained broadly uniform across strata, except in the top 10%, where 62% of individuals had SD(*P*_*i*_) ≤ 5 percentile points, consistent with the stronger inter-release correlation of SCZ scores.

To further characterise this pattern, we quantified rank stability using the invariability score *I*_*i*_, defined as the ratio of the mean to the standard deviation of individual percentile ranks across releases (Table S2; and Methods). Although *I*_*i*_ increased from lower to higher risk strata within each disorder, rising from 1.2 (IQR 0.9 to 1.5) in the BD bottom 10% to 8.5 (IQR 3.5 to 20.4) in the top 10%, from 1.1 (IQR 0.9 to 1.4) to 3.5 (IQR 2.2 to 6.3) in MDD, and from 1.4 (IQR 1.0 to 1.8) to 27.8 (IQR 12.4 to 57.5) in SCZ, this pattern reflects the mathematical scaling of the mean-to-SD ratio with percentile rank. It should therefore not be interpreted as evidence of reduced absolute rank variability at higher-risk levels. Consistent with this, rank variability and persistence were inversely related across all disorders and tolerance bands, with Spearman correlations ranging from −0.13 to −0.42 (Figure 2a), demonstrating that greater rank variability was associated with lower persistence relative to the most recent release.

**Figure 2:**
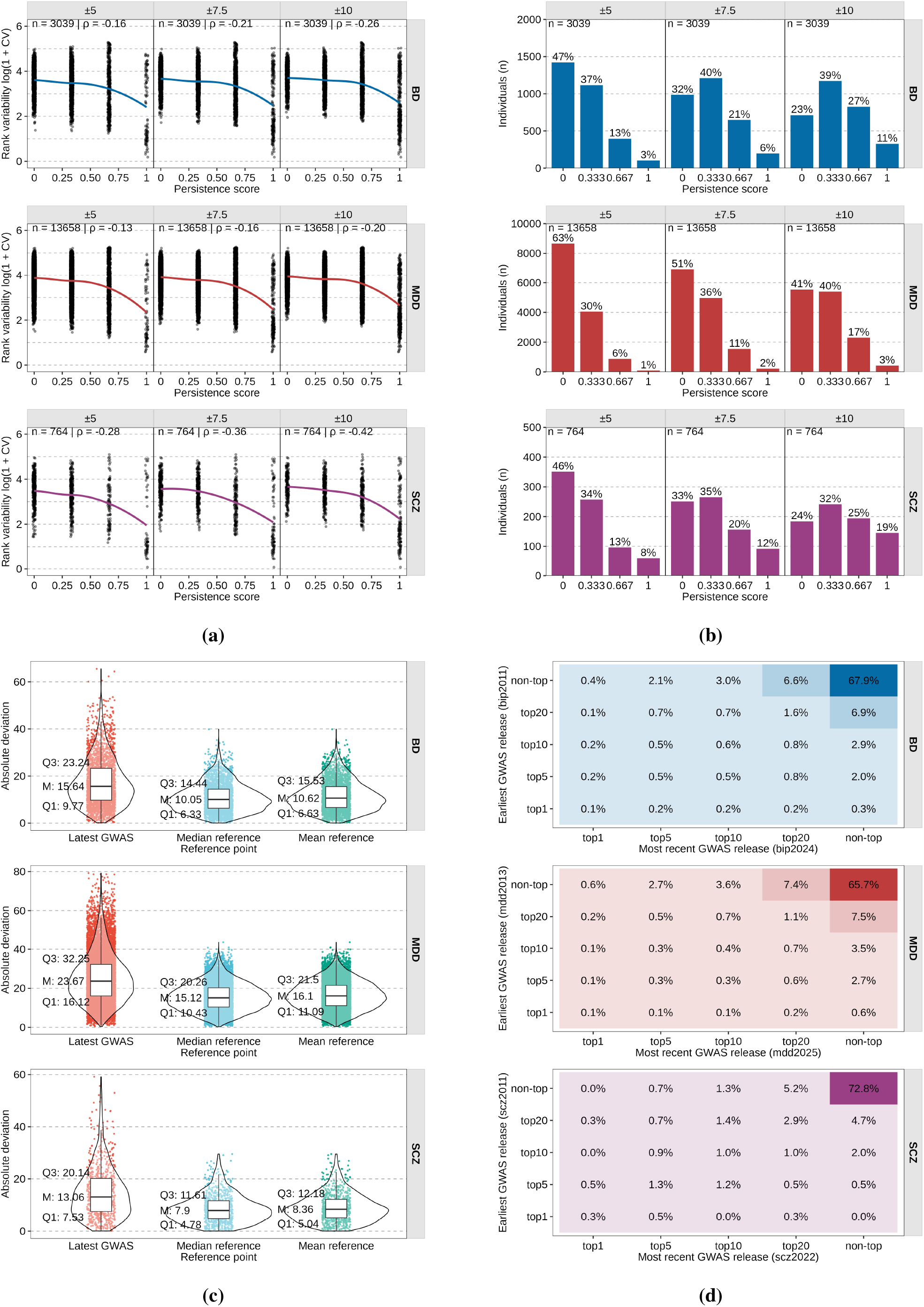
Structure and heterogeneity of within-individual PRS rank variability. **(a)** Scatter plots of rank variability against persistence score across three tolerance bands (±5, ±7.5, ±10), with Spearman correlations shown per disorder. **(b)** Bar plots showing the distribution of discrete persistence scores across three tolerance bands (±5, ±7.5, ±10). **(c)** Violin plots of absolute deviation (AD) from three reference points. Reclassification matrices showing the joint distribution of individuals across risk strata (top 1%, top 5%, top 10%, top 20%, non-top) between the earliest and most recent GWAS release.

To translate these findings into terms directly relevant to risk stratification, we quantified persistence within tolerance bands centred on the most recent release rank. Discrete persistence scores across tolerance bands of ±5, ±7.5, and ±10 percentile points (Methods) indicated that complete rank persistence was rare across all disorders (Table S3; Figure 2b). Within a ±5 percentile-point tolerance window, 47% of BD, 63% of MDD, and 46% of SCZ individuals fell outside the band at all prior releases, whereas complete rank persistence was observed for only 3%, 1%, and 8% of individuals, respectively. We next quantified absolute deviations relative to three reference points: the most recent release, the within-individual median, and the within-individual mean (Figure 2c). Median absolute deviations relative to the most recent release were 15.64 percentile points in BD, 23.67 in MDD, and 13.06 in SCZ. Re-anchoring to the within-individual median reduced this variability, though it did not eliminate it, to 10.05, 15.12, and 7.90 percentile points, respectively. Similar reductions were observed when anchoring to the within-individual mean, yielding deviations of 10.62, 16.10, and 8.36 percentile points, consistent with part of the observed variability reflecting fluctuations around an individual-specific central rank position. Nonetheless, substantial rank variability persisted regardless of the reference point used, motivating direct evaluation of whether individuals identified as high-risk retained that classification across successive releases.

### Retention in high-risk strata is variable across disorders and largely accounted for by inter-release correlation

Finally, we asked whether individuals identified as high risk under one release retained that designation as SNP effect-size estimates were iteratively updated across successive GWAS releases. We quantified observed retention at the top 10%, 5%, and 1% thresholds across successive releases and compared it with expected retention under a bivariate normal model parameterised by the observed inter-release correlation (Table S1 and S4; Figure 2d; and Methods). Two clear patterns emerged. First, retention varied across disorders and thresholds. For BD, top 10% retention ranged from 0.38 during the 2011-2019 transition, consistent with both the low inter-release correlation and the limited statistical power of the 2011 release, to 0.67 during the 2021-2024 transition, suggesting that retention is primarily governed by inter-release correlation rather than a stable individual property. For MDD, top-stratum retention was the lowest observed across disorders, ranging from 0.22 to 0.29, consistent with comparatively weaker inter-release correlations. For SCZ, top 10% retention ranged from 0.70 to 0.75 across the first two transitions, consistent with stronger inter-release correlation. Transitions into high-risk strata from outside were also common across all disorders (Figure 2d), with individuals originally outside the top stratum comprising 67.4%, 65.3%, and 72.8% of individuals in the most recent top stratum in BD, MDD, and SCZ, respectively, underscoring the bidirectional nature of reclassification at every threshold. Second, the retention gap between observed and expected values was small in BD and MDD, typically ranging from −0.05 to 0.10, but reached 0.38 in SCZ. These small gaps in BD and MDD indicate that observed retention in those disorders is largely accounted for by inter-release correlation, whereas SCZ exhibits modest excess retention beyond what the correlation structure alone would predict. Together, these results indicate that high-risk classifications across successive GWAS releases are determined primarily by the inter-release correlation structure rather than by stable individual rank positions.

## Discussion

These findings extend recent evidence of PRS rank variability across psychiatric disorders^18,19,20,21,22^ by providing a quantitative characterisation that distinguishes reclassification expected from model improvement from additional variability arising from the structure of polygenic score updating, while demonstrating that reclassification carries a genuine directional signal aligned with the trajectory of genetic discovery.

The directional consistency of empirical reclassification with case-control status demonstrates that successive GWAS releases carry informative signals at the individual level, with cases preferentially reclassified into higher-risk strata and controls into lower-risk strata at each successive improvement, though this signal coexists with structural rank variability that exceeds what improved discrimination alone would predict. This directional pattern is consistent with progressive attenuation of winner’s curse inflation^28^ in previously reported loci across successive releases, incorporation of newly associated loci, and increasingly well-calibrated effect-size estimates as discovery sample sizes grow. Excess reclassification beyond the theoretical minimum of expected reclassification reflects the high-dimensional geometry of polygenic architecture, in which simultaneous revisions to many per-SNP effect-size estimates induce nonlinear rank variability across percentile space that depends on individual variant profiles. This process is related to, but distinct from, uncertainty in individual PRS estimates arising from finite GWAS sample sizes within a single release^29^, and thus reflects an inherent property of iterative polygenic score improvement. Prior simulation work has established that ranking precision is a direct function of variance explained by a polygenic index22. The present findings empirically extend this relationship: transitions associated with larger gains in *R*^2^ produced stronger directional NRI, while the ratio of empirical to theoretical reclassification remained broadly consistent across disorders and successive releases.

Disorder-specific patterns also reflect differences in polygenic architecture and discovery trajectory. Schizophrenia, characterised by a more concentrated polygenic architecture and higher 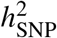, showed the smallest absolute deviations and strongest inter-release correlations, consistent with effect-size estimates approaching a more stable approximation of underlying liability as discovery matures. MDD, by contrast, with a more diffuse architecture and the largest expansions in discovery sample size, showed the greatest rank variability, consistent with recent evidence of lower rank stability and reduced specificity of MDD polygenic scores20.

The persistence and retention analyses place these findings in a clinical risk-stratification context. Complete rank persistence was rare across all disorders and tolerance bands, indicating that most individuals do not maintain stable positions relative to their most recent release rank, and that this variability is not concentrated within particular risk strata. Retention within high-risk strata varied across disorders and thresholds, with frequent movement both into and out of high-risk categories across releases. Retention gaps were small across most disorders and thresholds, indicating that preservation of high-risk classifications is governed largely by the inter-release correlation structure rather than by additional structure in individual rank trajectories. This extends prior observations of PRS rank variability in high-risk classifications19 by showing that much of this variability is, to a first approximation, explained by the inter-release correlation structure. Taken together, these results indicate that successive GWAS releases progressively refine individual PRS rankings in an informative direction, as evidenced by the consistent directionality of reclassification by case-control status. At the same time, excess reclassification beyond the theoretical minimum suggests that this improvement is accompanied by structural rank variability beyond that which improved discrimination alone would predict.

Several considerations limit the scope of the present findings. Analyses were based on four GWAS releases per disorder, which were sufficient to characterise the observed patterns but did not permit modelling of convergence trajectories or estimation of the discovery sample size at which individual PRS rankings may stabilise. All analyses were conducted in cohorts of European ancestry, and the extent to which these patterns generalise to non-European populations remains an important direction for future work^30^. The modest reductions in discriminative performance observed in the most recent BD and SCZ releases warrant particular attention for clinical implementation, as they resulted in negative NRI in European target samples. These performance reductions indicate that adoption of multi-ancestry releases may temporarily reverse the directional signal of reclassification in European cohorts, not because individuals’ underlying genetic liability has changed, but because cross-ancestry discovery data can attenuate transferability^15,31^. The use of a fixed SBayesRC parametrisation provides a conservative estimate of total rank variability. The present analyses also do not directly evaluate the relationship between rank variability and longitudinal clinical outcomes, which will be important for determining how reclassification magnitude translates into clinical decision-making settings.

These findings have implications for research and clinical settings in which PRS are used to stratify individuals by genetic risk. The directional consistency of reclassification with case-control status supports the use of successive GWAS releases to refine individual risk estimates, as each update carries a genuine signal that reflects improvements in discriminative performance. At the same time, excess reclassification beyond the theoretical minimum expected reclassification, limited rank persistence across releases, and variable retention of high-risk classifications indicate that individuals identified as high risk in one release may not retain that status as effect-size re-estimation progresses. This instability is particularly relevant for studies that recruit participants based on PRS rankings and for risk-stratification strategies that offer additional interventions to individuals at elevated genetic risk, both of which depend on risk classification remaining stable as discovery evolves. Collectively, these findings establish that individual PRS rankings are dynamic and shaped by progressive improvements in effect-size estimates. As psychiatric genomics advances, an explicit understanding of this dynamic will be essential for the nuanced interpretation of polygenic risk scores and for developing risk-stratification approaches that account for the evolving nature of genetic discovery.

## Methods

### Study cohorts

Genotype data were obtained from three Australian psychiatric research cohorts: the Genetics of Bipolar Disorder Study (GBP; *n* = 3,039) ^23^, the Australian Genetics of Depression Study (AGDS; *n* = 13,659) ^24^, and the ClozaGene Study (CLZ; *n* = 765) ^25^. A shared control sample (*n* = 7,320) for all three cohorts was drawn from the QSkin Sun and Health Study (QSkin2) ^26^, a population-based study of Queensland adults selected at random from the electoral roll (registration to vote is compulsory) and originally established to investigate melanoma and other skin cancers, and used here as an unselected population reference sample for PRS analyses. Study protocols were approved by the QIMR Berghofer Human Research Ethics Committee, and all participants provided written informed consent.

### Summary statistics

GWAS summary statistics were obtained from the PGC data portal in accordance with data use agreements. For each disorder, four GWAS releases were selected to represent successive stages of discovery within a common methodological lineage. For BD, releases from 2011^32^, 2019^1^, 2021^33^, and 2024^34^ were included; for MDD, releases from 2013^35^, 2018^2^, 2019^5^, and 2025^36^; and for SCZ, releases from 2011^37^, 2013^6^, 2014^38^, and 2022^3^. For releases in which Australian cohorts contributed to the discovery meta-analysis (MDD 2019 and 2025 and BD 2024, for which AGDS and GBP cohorts were included), leave-one-cohort-out summary statistics were used to exclude these samples, thereby reducing potential overlap between discovery and target datasets and limiting inflation in discriminative performance estimates.

### Polygenic risk score construction

Polygenic risk scores were constructed using SBayesRC implemented in GCTB v2.5.4^27^. Analyses were restricted to variants aligned to the GRCh37 reference genome. Standard variant-level quality control was applied prior to score construction, including exclusion of variants with low imputation quality (INFO < 0.8), low minor allele frequency (MAF < 0.01), strand ambiguity (A/T or C/G), or duplicate identifiers. For each GWAS, summary statistics were analysed in SBayesRC using a linkage disequilibrium (LD) reference panel derived from European-ancestry UK Biobank genotypes^39^, together with baselineLD v2.2 functional annotations^40^. SBayesRC parameters were held constant across releases, so differences among PRS reflected re-estimation of SNP effect sizes across successive GWAS iterations rather than changes in model specification. Individual-level PRS were then calculated from dosage data for each cohort using PLINK v2.00a6 LM^41^, with identical scoring procedures applied across cohorts to ensure comparability of the resulting risk estimates.

### Within-individual rank variability measures

Within-individual rank variability was quantified using two measures adapted from the *estar* R package^42^: percentile-rank *invariability* and *persistence* across four GWAS releases.

Invariability was defined as the inverse of the coefficient of variation (CV) of percentile ranks:

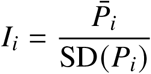

where 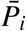 and SD(*P*_*i*_) denote the mean and standard deviation of individual *i*’s percentile ranks across releases. Higher *I*_*i*_ values indicate greater rank stability, whereas lower values indicate greater variability, implying substantial fluctuations in an individual’s position within the risk distribution across successive GWAS releases.

Persistence was defined as the proportion of prior releases in which percentile rank 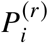 remained within a tolerance band δ of the final-release rank 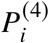:

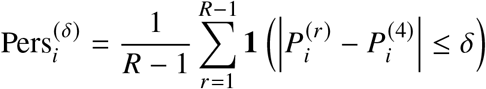

where *R* = 4 denotes the number of releases and δ ∈ {5, 7.5, 10} represents percentile-point tolerance bands. Values of 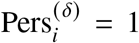 indicate complete rank persistence across all prior releases, whereas 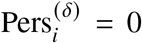 indicates no persistence relative to prior releases, suggesting that an individual’s most recent risk classification is unlikely to reflect classifications obtained under earlier GWAS releases.

### Reclassification analysis

Directional consistency between reclassification and case-control status was assessed using net reclassification improvement (NRI) for each consecutive GWAS release pair within each phenotype and cohort. Cases were defined as participants included in analyses for each target phenotype, whereas controls were drawn from the shared unselected reference sample used throughout the study.

For consecutive GWAS releases (*v, v* + 1), upward reclassification for individual *i* was defined as 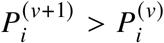, and downward reclassification as 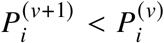. NRI was decomposed into case and control components:

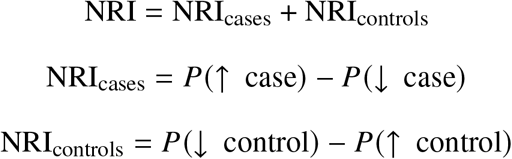

where *P*(↑ case) denotes the proportion of cases showing an increase in percentile rank from release *v* to *v* + 1, with analogous definitions for the remaining terms. Positive values of NRI_cases_ indicate preferential reclassification of cases into higher-risk strata, whereas positive values of NRI_controls_ indicate preferential reclassification of controls into lower-risk strata. A positive total NRI indicates improved discrimination between cases and controls.

### High-risk retention analysis

Retention in the upper tail was assessed using observed retention (Ret_obs_) and expected retention (Ret_exp_) for individuals initially above the 1%, 5%, and 10% thresholds across consecutive GWAS release pairs. Ret_obs_ was defined as the proportion of individuals above a threshold at release *v* who remained above that threshold at *v* + 1. Ret_exp_ was derived under a bivariate normal model incorporating the Pearson correlation ρ between consecutive PRS scores:

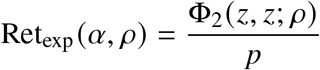

where *p* = (100 − *α*)/100, *z* = Φ^−1^(1 − *p*), and Φ_2_(·, ·; *ρ*) denotes the bivariate standard normal cumulative distribution function, computed using the *mvtnorm* R package^43^. The retention gap, defined as

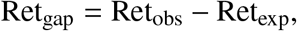

quantifies excess retention beyond that expected from inter-release correlation.

## Data Availability

GWAS summary statistics used in this study are publicly available from the Psychiatric Genomics Consortium data portal (https://pgc.unc.edu/for-researchers/download-results). All data produced in the present work are contained in the manuscript.

## Data availability

GWAS summary statistics used in this study are publicly available from the Psychiatric Genomics Consortium data portal (https://pgc.unc.edu/for-researchers/download-results/).

## Code availability

All analyses were performed using publicly available software. SBayesRC (https://github.com/zhilizheng/SBayesRC) was used to re-weight SNP effect sizes for polygenic risk prediction. Individual-level polygenic risk scores were calculated using PLINK v2.00a6 LM (https://www.cog-genomics.org/plink/2.0/). Statistical analyses and visualisations were performed in R (https://www.r-project.org/), using the packages *ggplot2, patchwork, mvtnorm* v1.x.x and *estar*.

## Acknowledgements

The authors thank all participants from the QSkin Sun and Health Study, the Genetics of Bipolar Disorder Study, the Australian Genetics of Depression Study, and the ClozaGene Study for giving their time and support for this project. For the AGDS, the authors also wish to thank all the people who helped in the conception, implementation, beta testing, media campaign and data cleaning. The authors would specifically like to acknowledge K. Kendler, P. Sullivan, A. McIntosh and C. Lewis for input on the questionnaire; L. Nunn, M. Ferguson, L. Winkler and N. Garden for data and sample collection; J. Davies, L. Lowrey and V. Antonini for support with IT aspects; V. Morgan and K. Kirkby for help with the media campaign. The authors would also like to thank N. Wray for her helpful review and contributions to the manuscript. The QSkin Sun and Health Study was supported by grants from the National Health and Medical Research Council (NHMRC) of Australia (APP1073898; APP1063061; APP1185416; APP2030931). David Whiteman is supported by an NHMRC Investigator Grant (APP2026567). The authors acknowledge and thank M. Steffens for her generous donations in loving memory of J. Banks. Data collection was funded and data analysis was supported by the Australian National Health and Medical Research Council (No. APP1138514) to S.E.M. D.J.S. is supported by a National Health and Medical Research Council Investigator Grant (No. APP1194635). S.E.M. is supported by a National Health and Medical Research Council Investigator Grant (No. APP2025674). The AGDS was primarily funded by the Australian National Health and Medical Research Council (NHMRC) grant (No. APP1086683 to NGM). This work was further supported by the NHMRC (grant numbers 1145645, 1078901 and 1087889). NGM was supported by an NHMRC Investigator Grant (No. APP1172990). SEM is supported by an NHMRC Investigator Grant (No. APP2025674). The ClozaGene Study was supported by the Australian National Health and Medical Research Council (Project Grant APP1138514 to S.E.M.). S.E.M. is supported by Australian National Health and Medical Research Council Investigator Grant (No. APP2025674). DJS is supported by a UQ Amplify Professorial Research Fellowship. The funders had no role in the study design, data collection and analysis, decision to publish or preparation of the manuscript.

## Author information

### Authors and Affiliations

Brain and Mental Health, QIMR Berghofer Medical Research Institute, Brisbane, QLD, Australia Andreza Ferreira, Penelope A. Lind, Nicholas G. Martin & Sarah E. Medland

Academic Division, Education Portfolio, Queensland University of Technology, Brisbane, QLD, Australia

Andreza Ferreira, Hayley Moody & Richard C. Medland

School of Biomedical Sciences, Faculty of Health, Queensland University of Technology, Brisbane, QLD, Australia

Penelope A. Lind & Matthew H. Law

School of Biomedical Sciences, Faculty of Health, Medicine and Behavioural Sciences, The University of Queensland, Brisbane, QLD, Australia

Penelope A. Lind, Matthew H. Law & Dan J. Siskind

Brain and Mind Centre, University of Sydney, Camperdown, NSW, Australia

Ian B. Hickie

Population Health, QIMR Berghofer Medical Research Institute, Brisbane, QLD, Australia Catherine M. Olsen, David C. Whiteman & Matthew H. Law

Addiction and Mental Health Service, Metro South Health, Brisbane, QLD, Australia Dan J. Siskind

Physical and Mental Health Stream, Queensland Centre for Mental Health Research, Brisbane, QLD, Australia

Dan J. Siskind

School of Psychology and Counselling, Queensland University of Technology, Brisbane, QLD, Australia

Sarah E. Medland

School of Psychology, The University of Queensland, Brisbane, QLD, Australia Sarah E. Medland

### Contributions

A.F. conducted the central analyses and wrote the manuscript. I.B.H., C.M.O., D.C.W., M.H.L., D.J.S., N.G.M., P.A.L. and S.E.M. collected samples, conducted phenotyping and genotyping, and led the cohorts whose data were included in the study. P.A.L., H.M., R.C.M. and S.E.M. worked closely with A.F. on the design and conduct of the study. All authors contributed to critical revisions of the manuscript and approved the final version for submission.

## Ethics declarations

### Competing interests

Coauthor DS serves on the Viatris Australian Clozapine Quality Advisory Committee and has received speaker and/or consultancy fees from Servier, Otsuka, Viatris and Lundbeck. Coauthor IBH is the Co-Director of Health and Policy at the Brain and Mind Centre (BMC) University of Sydney. The BMC operates an early-intervention youth service at Camperdown under contract to Headspace. He is the Chief Scientific Advisor to, and a 3.2% equity shareholder in, InnoWell Pty Ltd. InnoWell was formed by the University of Sydney (45% equity) and PwC (Australia; 45% equity) to deliver the $30 M Australian Government-funded Project Synergy (2017-20; a three-year program for the transformation of mental health services) and to lead transformation of mental health services internationally through the use of innovative technologies. The other coauthors declare that there are no conflicts of interest with respect to the research, authorship and/or publication of this article.

---

**Table S1:** Pairwise Spearman rank correlations of individual PRS percentile ranks across GWAS releases.

| Release | bip2011 | bip2019 | bip2021 | bip2024 | mdd2013 | mdd2018 | mdd2019 | mdd2025 | scz2011 | scz2013 | scz2014 | scz2022 |
| --- | --- | --- | --- | --- | --- | --- | --- | --- | --- | --- | --- | --- |
| bip2011 | 1.00 | 0.56 | 0.45 | 0.41 | * | * | * | * | * | * | * | * |
| bip2019 |  | 1.00 | 0.79 | 0.71 | * | * | * | * | * | * | * | * |
| bip2021 |  |  | 1.00 | 0.88 | * | * | * | * | * | * | * | * |
| bip2024 |  |  |  | 1.00 | * | * | * | * | * | * | * | * |
|  | * | * | * | * | * | * | * | * | * | * | * | * |
| mdd2013 | * | * | * | * | 1.00 | 0.29 | 0.14 | 0.19 | * | * | * | * |
| mdd2018 | * | * | * | * |  | 1.00 | 0.42 | 0.54 | * | * | * | * |
| mdd2019 | * | * | * | * |  |  | 1.00 | 0.42 | * | * | * | * |
| mdd2025 | * | * | * | * |  |  |  | 1.00 | * | * | * | * |
|  | * | * | * | * | * | * | * | * | * | * | * | * |
| scz2011 | * | * | * | * | * | * | * | * | 1.00 | 0.86 | 0.73 | 0.68 |
| scz2013 | * | * | * | * | * | * | * | * |  | 1.00 | 0.79 | 0.74 |
| scz2014 | * | * | * | * | * | * | * | * |  |  | 1.00 | 0.88 |
| scz2022 | * | * | * | * | * | * | * | * |  |  |  | 1.00 |

BD, bipolar disorder; MDD, major depressive disorder; SCZ, schizophrenia; $\rho_S$ , Spearman rank correlation of individual PRS percentile ranks between two GWAS releases; higher values indicate stronger preservation of individual rank ordering across releases; asterisk (\*) indicates cross-disorder pairs, not computed; lower triangle left blank.

**Table S2:** Within-individual rank variability metrics: invariability score (*I*_*i*_) by risk stratum.

| Disorder | Risk stratum | *n* | SD < 5 | SD 5–10 | SD 10–20 | SD ≥ 20 | *I* median [IQR] |
| --- | --- | --- | --- | --- | --- | --- | --- |
| BD | Overall | 3,039 | 266 (9%) | 636 (21%) | 1,330 (44%) | 807 (27%) | 2.9 [1.8–5.2] |
|  | Bottom 10% (<10) | 304 | 80 (26%) | 61 (20%) | 74 (24%) | 89 (29%) | 1.2 [0.9–1.5] |
|  | Lower-middle (10–49) | 1,215 | 45 (4%) | 253 (21%) | 608 (50%) | 309 (25%) | 2.3 [1.6–3.2] |
|  | Upper-middle (50–89) | 1,216 | 59 (5%) | 259 (21%) | 574 (47%) | 324 (27%) | 4.4 [2.7–7.2] |
|  | Top 10% (≥90) | 304 | 82 (27%) | 63 (21%) | 74 (24%) | 85 (28%) | 8.5 [3.5–20.4] |
| MDD | Overall | 13,658 | 315 (2%) | 1,166 (9%) | 4,569 (33%) | 7,608 (56%) | 2.1 [1.5–3.3] |
|  | Bottom 10% (<10) | 1,366 | 79 (6%) | 148 (11%) | 382 (28%) | 757 (55%) | 1.1 [0.9–1.4] |
|  | Lower-middle (10–49) | 5,463 | 78 (1%) | 419 (8%) | 1,932 (35%) | 3,034 (56%) | 1.8 [1.4–2.5] |
|  | Upper-middle (50–89) | 5,463 | 84 (2%) | 454 (8%) | 1,860 (34%) | 3,065 (56%) | 2.7 [1.8–4.4] |
|  | Top 10% (≥90) | 1,366 | 74 (5%) | 145 (11%) | 395 (29%) | 752 (55%) | 3.5 [2.2–6.3] |
| SCZ | Overall | 764 | 132 (17%) | 199 (26%) | 340 (45%) | 93 (12%) | 3.5 [2.0–7.0] |
|  | Bottom 10% (<10) | 77 | 26 (34%) | 19 (25%) | 17 (22%) | 15 (19%) | 1.4 [1.0–1.8] |
|  | Lower-middle (10–49) | 305 | 17 (6%) | 83 (27%) | 171 (56%) | 34 (11%) | 2.5 [1.8–3.5] |
|  | Upper-middle (50–89) | 305 | 41 (13%) | 82 (27%) | 138 (45%) | 44 (14%) | 5.4 [3.4–9.8] |
|  | Top 10% (≥90) | 77 | 48 (62%) | 15 (19%) | 14 (18%) | 0 (0%) | 27.8 [12.4–57.5] |

BD, bipolar disorder; MDD, major depressive disorder; SCZ, schizophrenia. *n*, number of individuals in each stratum; SD, standard deviation of individual percentile ranks across four GWAS releases, in percentile points; counts reported as *n* (%) within each SD band; *I*, ratio of mean to standard deviation of individual percentile ranks across releases; higher values indicate greater rank stability.

**Table S3:** Within-individual rank variability metrics: persistence score 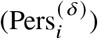 by tolerance band and risk stratum.

| Disorder | Risk stratum | $\delta$ | Pers = 0 | Pers = 0.33 | Pers = 0.67 | Pers = 1.00 |
| --- | --- | --- | --- | --- | --- | --- |
| BD | Overall | $\pm 5$ | 1,423 (47%) | 1,117 (37%) | 396 (13%) | 103 (3%) |
| | | $\pm 7.5$ | 986 (32%) | 1,209 (40%) | 647 (21%) | 197 (6%) |
| | | $\pm 10$ | 713 (23%) | 1,171 (39%) | 827 (27%) | 328 (11%) |
| | Top 10% | $\pm 5$ | 62 (20%) | 111 (37%) | 88 (29%) | 43 (14%) |
| | | $\pm 7.5$ | 42 (14%) | 88 (29%) | 105 (35%) | 69 (23%) |
| | | $\pm 10$ | 31 (10%) | 79 (26%) | 108 (36%) | 86 (28%) |
| MDD | Overall | $\pm 5$ | 8,660 (63%) | 4,045 (30%) | 867 (6%) | 86 (1%) |
| | | $\pm 7.5$ | 6,933 (51%) | 4,970 (36%) | 1,542 (11%) | 213 (2%) |
| | | $\pm 10$ | 5,536 (41%) | 5,402 (40%) | 2,297 (17%) | 423 (3%) |
| | Top 10% | $\pm 5$ | 666 (49%) | 471 (34%) | 200 (15%) | 29 (2%) |
| | | $\pm 7.5$ | 517 (38%) | 502 (37%) | 289 (21%) | 58 (4%) |
| | | $\pm 10$ | 425 (31%) | 519 (38%) | 335 (25%) | 87 (6%) |
| SCZ | Overall | $\pm 5$ | 351 (46%) | 257 (34%) | 96 (13%) | 60 (8%) |
| | | $\pm 7.5$ | 251 (33%) | 265 (35%) | 156 (20%) | 92 (12%) |
| | | $\pm 10$ | 184 (24%) | 241 (32%) | 194 (25%) | 145 (19%) |
| | Top 10% | $\pm 5$ | 15 (19%) | 17 (22%) | 12 (16%) | 33 (43%) |
| | | $\pm 7.5$ | 9 (12%) | 10 (13%) | 19 (25%) | 39 (51%) |
| | | $\pm 10$ | 4 (5%) | 11 (14%) | 16 (21%) | 46 (60%) |

BD, bipolar disorder; MDD, major depressive disorder; SCZ, schizophrenia. $\delta$ , tolerance band in percentile points, tested at $\pm 5$ , $\pm 7.5$ , and $\pm 10$ ; $\text{Pers}_i^{(\delta)}$ , proportion of the three prior releases in which an individual's percentile rank remained within $\delta$ percentile points of the most recent release; Pers = 0, absence of rank persistence at all prior releases; Pers = 1.00, complete rank persistence within the tolerance band across all prior releases.

**Table S4:** Observed and expected retention in high-risk strata across successive GWAS releases.

| Disorder | Transition | Risk stratum | $n_{\text{stratum}}$ | $\rho_{\text{PRS}}$ | $\text{Ret}_{\text{obs}}$ | $\text{Ret}_{\text{exp}}$ | $\text{Ret}_{\text{gap}}$ |
| --- | --- | --- | --- | --- | --- | --- | --- |
| BD | 2011 → 2019 | Top 10% | 304 | 0.56 | 0.38 | 0.36 | +0.02 |
|  |  | Top 5% | 152 | 0.56 | 0.29 | 0.28 | +0.01 |
|  |  | Top 1% | 31 | 0.56 | 0.23 | 0.16 | +0.07 |
| | 2019 → 2021 | Top 10% | 304 | 0.79 | 0.58 | 0.55 | +0.03 |
|  |  | Top 5% | 152 | 0.79 | 0.57 | 0.48 | +0.09 |
|  |  | Top 1% | 31 | 0.79 | 0.45 | 0.36 | +0.09 |
| | 2021 → 2024 | Top 10% | 304 | 0.88 | 0.67 | 0.67 | +0.01 |
|  |  | Top 5% | 152 | 0.88 | 0.65 | 0.61 | +0.04 |
|  |  | Top 1% | 31 | 0.88 | 0.61 | 0.51 | +0.10 |
| MDD | 2013 → 2018 | Top 10% | 1,366 | 0.29 | 0.22 | 0.21 | +0.01 |
|  |  | Top 5% | 683 | 0.29 | 0.13 | 0.14 | -0.00<sup>a</sup> |
|  |  | Top 1% | 137 | 0.29 | 0.07 | 0.05 | +0.02 |
| | 2018 → 2019 | Top 10% | 1,366 | 0.42 | 0.29 | 0.28 | +0.01 |
|  |  | Top 5% | 683 | 0.42 | 0.20 | 0.20 | +0.00 |
|  |  | Top 1% | 137 | 0.42 | 0.07 | 0.09 | -0.02<sup>a</sup> |
| | 2019 → 2025 | Top 10% | 1,366 | 0.42 | 0.27 | 0.28 | -0.01<sup>a</sup> |
|  |  | Top 5% | 683 | 0.42 | 0.21 | 0.20 | +0.01 |
|  |  | Top 1% | 137 | 0.42 | 0.04 | 0.09 | -0.05<sup>a</sup> |
| SCZ | 2011 → 2013 | Top 10% | 77 | 0.86 | 0.75 | 0.63 | +0.12 |
|  |  | Top 5% | 39 | 0.86 | 0.90 | 0.57 | +0.33 |
|  |  | Top 1% | 8 | 0.86 | 0.50 | 0.46 | +0.04 |
| | 2013 → 2014 | Top 10% | 77 | 0.79 | 0.70 | 0.55 | +0.15 |
|  |  | Top 5% | 39 | 0.79 | 0.77 | 0.48 | +0.29 |
|  |  | Top 1% | 8 | 0.79 | 0.38 | 0.36 | +0.02 |
| | 2014 → 2022 | Top 10% | 77 | 0.88 | 0.73 | 0.66 | +0.07 |
|  |  | Top 5% | 39 | 0.88 | 0.62 | 0.60 | +0.01 |
|  |  | Top 1% | 8 | 0.88 | 0.88 | 0.50 | +0.38 |

BD, bipolar disorder; MDD, major depressive disorder; SCZ, schizophrenia; $n_{\text{stratum}}$ , number of individuals above threshold at the earlier release; $\rho_{\text{PRS}}$ , Pearson correlation of polygenic risk score percentile ranks between releases; $\text{Ret}_{\text{obs}}$ , proportion of individuals above threshold at the earlier release who remain above threshold at the later release; $\text{Ret}_{\text{exp}}$ , expected retention under a bivariate normal model with inter-release correlation $\rho_{\text{PRS}}$ ; $\text{Ret}_{\text{gap}} = \text{Ret}_{\text{obs}} - \text{Ret}_{\text{exp}}$ . <sup>a</sup> Negative $\text{Ret}_{\text{gap}}$ indicates that observed retention was lower than expected under the bivariate normal model.

## Notes

### Funding Statement

QSkin: The QSkin Sun and Health Study was supported by grants from the National Health and Medical Research Council (NHMRC) of Australia (APP1073898; APP1063061; APP1185416; APP2030931). David Whiteman is supported by an NHMRC Investigator Grant (APP2026567). GBP: We acknowledge and thank M. Steffens for her generous donations in loving memory of J. Banks. Data collection was funded and data analysis was supported by the Australian National Health and Medical Research Council (No. APP1138514) to S.E.M. D.J.S. is supported by a National Health and Medical Research Council Investigator Grant (No. APP1194635). S.E.M. is supported by a National Health and Medical Research Council Investigator Grant (No. APP2025674). AGDS: The AGDS was primarily funded by the Australian National Health and Medical Research Council (NHMRC) grant (No. APP1086683 to NGM). This work was further supported by the NHMRC (grant numbers 1145645, 1078901 and 1087889). NGM was supported by an NHMRC Investigator Grant (No. APP1172990). SEM is supported by an NHMRC Investigator Grant (No. APP2025674). ClozaGene: This work was supported by the Australian National Health and Medical Research Council (Project Grant APP1138514 to S.E.M.). S.E.M. is supported by Australian National Health and Medical Research Council Investigator Grant (No. APP2025674). DJS is supported by a UQ Amplify Professorial Research Fellowship.

### Author Declarations

Ethics committee of QIMR Berghofer gave ethical approval for this work.

